# OmicFormer: a statistical priors–informed transformer for accurate and generalizable omics prediction of diseases and complex traits

**DOI:** 10.64898/2026.07.06.26357359

**Authors:** Hanlin Jiang, Chang Yang, Menhan Qin, Jia You, Jianfeng Feng, Jin-Tai Yu, Wei Cheng, Weikang Gong

## Abstract

Precision medicine faces a critical challenge in translating high-dimensional omics data into robust disease predictions across diverse populations. Current approaches often fail under distribution shifts, partly due to their inability to encode complex biological feature dependencies. We present OmicFormer, a Transformer-based architecture that embeds two complementary statistical priors, i.e., feature–label associations and feature–feature dependencies, directly into its representation learning. This design captures local and long-range omic interactions often missed by conventional methods. Analyzing 500,000 UK Biobank participants, OmicFormer significantly outperforms strong baselines across 450 disease and 900 trait prediction tasks, with substantial gains spanning diverse metabolic, neurological, cardiovascular, and gastrointestinal conditions, alongside enhanced prediction of circulating metabolites, bone density traits, and retinal imaging biomarkers. Crucially, OmicFormer demonstrates robust generalization, achieving a substantial improvement over tree-based methods in an independent proteomics cohort across 19 diseases (GNPC, N=7,289), and outperforming tree-based models across 50 multi-site neuroimaging sites (N=4,728) for autism and schizophrenia classification. By explicitly embedding statistical structure, OmicFormer provides an interpretable and generalizable foundation for omics-based precision medicine.

## Introduction

Over the past decade, the rapid advancement of precision medicine has been accompanied by an unprecedented expansion of multimodal biomedical omics data, creating new opportunities for disease diagnosis, risk prediction, and biomarker discovery^1,2^. Large-scale cohort resources such as the UK Biobank (UKB) systematically integrate diverse omic modalities, including clinical phenotypes, genomics, proteomics, metabolomics, and imaging-derived phenotypes^3,4^, providing a strong empirical foundation for dissecting molecular mechanisms and phenotypic heterogeneity in complex traits and diseases. Nevertheless, effectively extracting actionable knowledge from these high-dimensional, nonlinear, heterogeneous, and often incomplete omic datasets remains a central challenge for AI-driven healthcare^5,6^.

Omic data are typically structured as tabular datasets, where rows correspond to individuals and columns represent features. Despite their ubiquity, such tabular representations pose inherent analytical difficulties: feature spaces are large (often hundreds to thousands of dimensions), interdependencies among variables are complex and often nonlinear, and clinically relevant signals are usually distributed across many weakly predictive features rather than concentrated in a few strong predictors.

Existing modeling approaches, including simple linear regression, LASSO, and in particular, tree-based methods such as LightGBM^7^, have long served as default choices for omics prediction. However, these approaches suffer from fundamental limitations in high-dimensional omics settings: linear models assume additive effects and cannot capture complex nonlinear interactions, while tree-based methods operate through greedy, axis-aligned splitting that treats features independently and cannot model global interactions. Moreover, both of them lack mechanisms to incorporate prior biological knowledge (e.g., biological networks); and their performance degrades substantially under distribution shifts across cohorts, limiting real-world deployment^8^.

Deep learning methods have been introduced for tabular data analysis to address some of these limitations, yet their adoption in omics remains limited^9^. Architectures such as TabNet^10^ and TabMap^11^ can improve predictive performance but face persistent challenges: they introduce substantial complexity without consistently outperforming well-tuned tree-based baselines in omic datasets^9^. And most critically, existing approaches do not explicitly encode the statistical dependencies that are inherent to biological systems, such as protein–protein interaction networks, metabolic pathway correlations, or imaging-derived structural covariance, into their representation learning pipelines. This omission means that even deep models may fail to leverage the full structure of omics data, instead treating features as independent entities until interactions are (implicitly and expensively) learned from data.

To address these challenges, we introduce OmicFormer, a framework that explicitly injects statistical structure into Transformer-based representation learning for omics tabular data. Unlike previous approaches that rely on implicit interaction learning, OmicFormer integrates correlation-guided inductive biases directly into the input representation through two complementary channels. The first orders features by their correlation with the task labels, placing predictive features in close proximity to enable more effective local pattern extraction. The second derives a structured ordering from the feature–feature correlation matrix via Gromov–Wasserstein optimal transport alignment^12^, explicitly encoding intrinsic biological dependencies, such as co-expressed protein modules or correlated brain regions, into the sequence layout. On top of these dual-channel inputs, a multi-scale patch embedding module extracts patterns across varying receptive fields, while a Transformer encoder models long-range dependencies across the entire feature sequence. This design directly addresses the inability of tree-based methods to model global interactions and the tendency of standard deep learning approaches to treat features as independent until trained.

To further improve generalization, we incorporate multi-task learning (MTL) during training. By jointly optimizing related targets within a shared representation space, MTL enables cross-task knowledge transfer and acts as an implicit regularizer^13,14^, particularly valuable for rare diseases. In disease classification, auxiliary tasks are selected based on comorbidity structures^15,16^; for continuous phenotypes, auxiliary targets are chosen according to label correlations^17,18^.

OmicFormer is broadly applicable to diverse omics datasets. Using data from approximately 500,000 UK Biobank participants, we demonstrate its performance in predicting 15-year disease risks for 450 diseases and estimating 900 health-related traits across plasma proteomics, metabolomics and brain imaging phenotypes. Crucially, in external validation studies on independent cohorts for proteomics-based disease prediction (GNPC^19^, N=7,289) and multi-site neuroimaging cohorts for autism^20^ and schizophrenia prediction^21^ (N=4,728, 50 different sites), OmicFormer is compared with strong baselines under realistic distribution shifts, addressing the generalization gap that limits tree-based methods in real-world deployment and underscoring its potential for multi-center precision medicine^3,22–24^.

## Results

### Overview of cohorts

We primarily evaluate OmicFormer on the UK Biobank (UKB)^3^, leveraging three modalities: (1) Plasma proteomics from the UKB Pharma Proteomics Project (UKB-PPP; Olink Explore 3072; 2,920 proteins; ∼54,000 samples); (2) Blood metabolomics profiled using the Nightingale Health NMR platform; (3) Brain imaging-derived phenotypes (IDPs) computed from structural, diffusion, and functional MRI pipelines. To assess robustness under cross-cohort distribution shift, we further perform external validation for proteomics on the Global Neurodegeneration Proteomics Consortium (GNPC)^19^, restricting evaluation to the 2,137 proteins shared between UKB-PPP and GNPC and testing models trained on UKB directly on GNPC (no fine-tuning). For neuroimaging data, we additionally validate OmicFormer on two independent multi-site cohorts, ABIDE^20^ for autism prediction (34 sites; resting-state fMRI functional connectivities) and ZIB^25^ for schizophrenia prediction (16 sites, structural MRI-based IDPs), which provide heterogeneous acquisition conditions for evaluating cross-site reproducibility.

### OmicFormer enables accurate disease risk prediction

Using omics data from the UK Biobank, we systematically evaluated OmicFormer against widely used baselines for future disease risk prediction across proteomics, metabolomics, and brain imaging-derived phenotypes (IDPs). The compared methods included LightGBM, 1D-CNN, TabMap, and TabNet (Fig. 2a). Detailed disease-level results for the three modalities are provided in Supplementary Table 1–3. While OmicFormer achieved highly significant average improvements across all modalities, a disease-level analysis further revealed that these gains translated into substantial improvements for a wide range of individual diseases (Fig. 2b).

**Figure 1.**
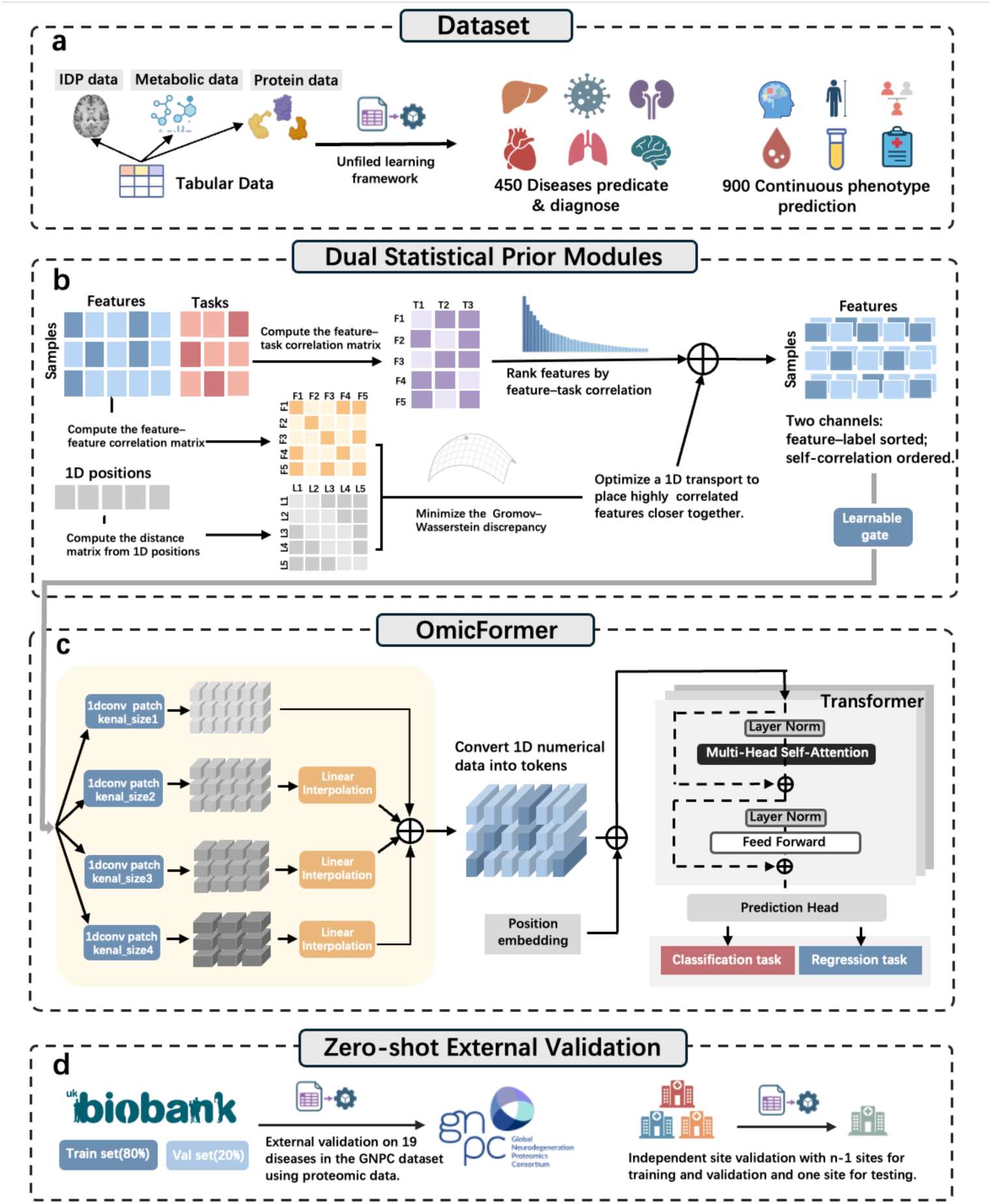
Overview of the OmicFormer. a. Multimodal omics data (proteomics, metabolomics, imaging-derived phenotypes) from UK Biobank are unified under a tabular learning framework for disease prediction (450 diseases) and continuous trait regression (900 quantitative traits). b. A dual-channel semantic ordering strategy ranks features by feature–task correlation, then applies Gromov–Wasserstein optimal transport to align feature–feature correlations with 1D positional distance, placing correlated features nearby while preserving tabular format. c. Multi-scale 1D convolutional patch embedding layers extract hierarchical local patterns; tokens are processed by a Transformer encoder with multi-head self-attention and feed-forward layers, followed by a task-specific prediction head. d. Generalization is evaluated via external validation on the GNPC proteomics cohort and leave-one-site-out validation in multi-center datasets.

**Figure 2.**
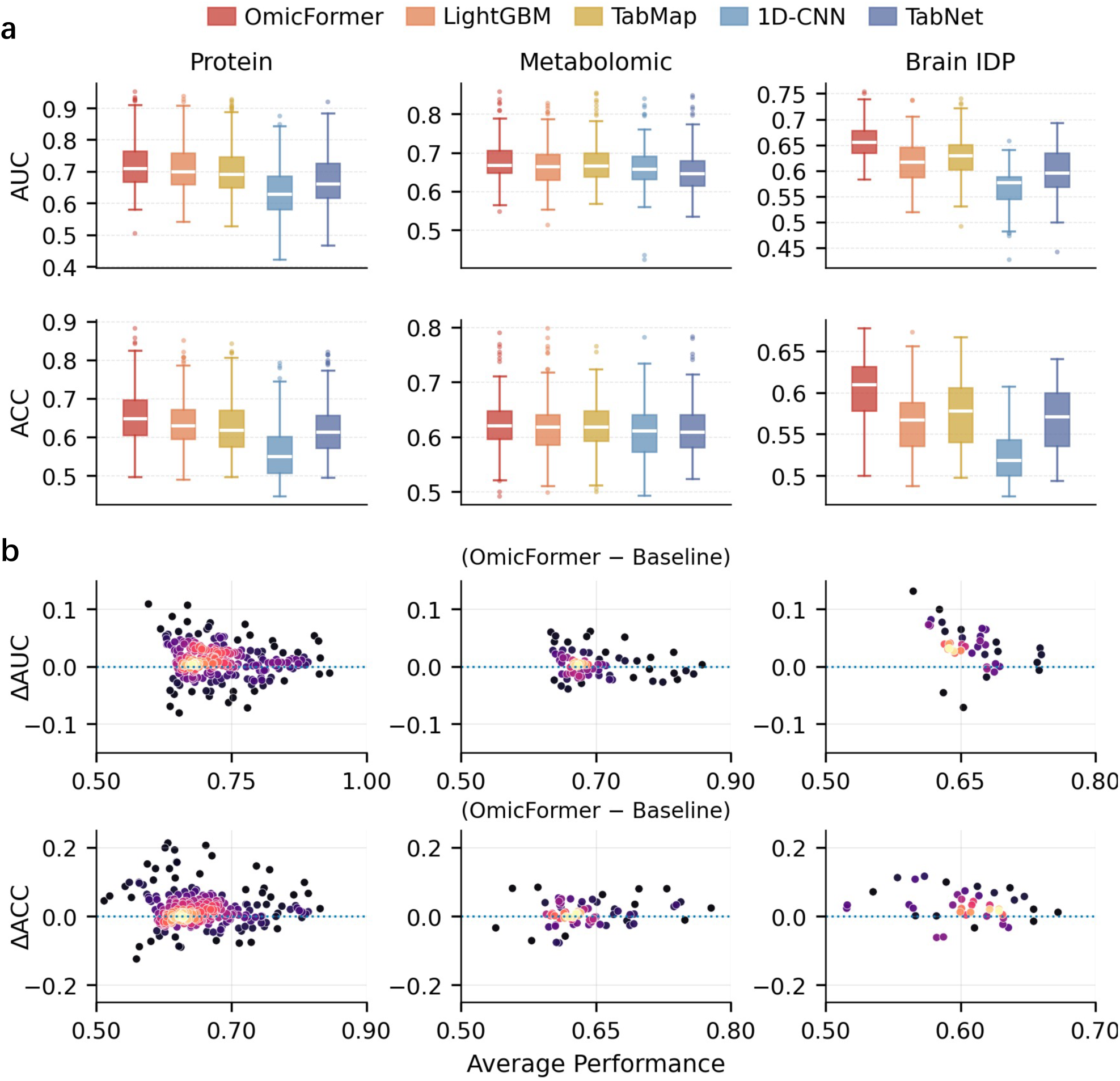
OmicFormer performance and improvement across omics datasets for future disease risk prediction. a. Predictive performance (AUC, top; ACC, bottom) across proteomics, metabolomics, and imaging-derived phenotypes (IDPs). OmicFormer consistently outperforms all baseline methods (LightGBM, TabMap, TabNet, and 1D-CNN), achieving higher mean performance and lower variance across 450 diseases. b. Scatter plots showing per-disease performance differences (Δ = OmicFormer − baseline) between OmicFormer and the strongest competing baseline for each modality. AUC differences are shown in the top row and ACC differences in the bottom row, with positive values indicating improvement.

In proteomics, OmicFormer achieved a mean AUC of 0.724, significantly exceeding the strongest baseline (LightGBM: 0.714) with a 3.53% relative improvement (p = 1.36 × 10^-9^). Mean ACC increased from 0.636 to 0.656, corresponding to a 5.44% relative improvement (p = 7.01 × 10⁻⁸). OmicFormer also showed larger margins over other competitive baselines, including TabMap (6.02% AUC relative improvement; p = 1.38 × 10⁻¹⁹) and TabNet (15.35% AUC relative improvement; p = 7.08 × 10⁻⁴⁵), indicating broad improvements beyond any single comparator. At the individual disease level, these highly significant average gains were underpinned by pronounced improvements for numerous conditions. Representative AUC gains included diabetic neuropathy (0.911 to 0.954; +47.81%), vascular dementia (0.647 to 0.777; +36.76%), and hypertensive renal disease (0.924 to 0.951; +35.74%).

In metabolomics, OmicFormer achieved a mean AUC of 0.681, which is similar to TabMap, the strongest baseline in this modality (0.678; 0.46% relative improvement, p = 2.78 × 10⁻¹). For ACC, OmicFormer reached 0.626, compared with 0.620 for TabMap (0.98% relative improvement, p = 1.07 × 10⁻¹). Despite the similar average performance, at the individual disease level, OmicFormer delivered substantial gains for a substantial subset of diseases. For example, OmicFormer achieved notable AUC gains for hemiplegia (0.538 to 0.658; +25.93%), cerebral palsy and other paralytic syndromes (0.568 to 0.655; +20.12%), and duodenal ulcer (0.667 to 0.733; +19.72%).

In brain IDPs, OmicFormer achieved a mean AUC of 0.659, outperforming TabMap, the strongest baseline in this modality, by 5.11% relative improvement (0.659 vs. 0.627, p = 1.11 × 10⁻⁸). For ACC, OmicFormer reached 0.605, exceeding TabMap (0.573) by 5.64% relative improvement (p = 2.25 × 10⁻⁷). At the individual disease level, the gains were again widespread and substantial, with representative AUC improvements including type 2 diabetes (0.691 to 0.745; +17.54%), disorders of gallbladder, biliary tract and pancreas (0.589 to 0.651; +15.05%), and acute renal failure (0.642 to 0.693; +14.40%).

Given the large number of heterogeneous diseases, achieving statistically significant average improvements represents a stringent benchmark. The diversity in prediction difficulty, biological mechanisms, and signal-to-noise ratios makes even modest average gains nontrivial. In this context, OmicFormer’s consistent and highly significant average improvements, together with its widespread gains at the individual phenotype level, underscore the robustness and generalizability of the proposed approach across a broad spectrum of disease prediction tasks.

### Stratified performance of OmicFormer across disease chapters

In proteomics-to-disease prediction, stratifying outcomes into broad disease chapters revealed a clear and clinically intuitive performance pattern. OmicFormer consistently outperformed all baselines at the chapter level, with chapter-wise mean AUCs spanning 0.676–0.782. The most pronounced gains (visible across every baseline comparator) occurred in chapters where baselines were notably weaker, such as gastrointestinal (ΔAUC = +0.022 vs LightGBM; +6.42% relative improvement; p = 2.39 × 10⁻³; n = 32) and infectious (+0.019; +6.18%; p = 2.52 × 10⁻⁴; n = 18). By contrast, chapters with uniformly strong performance across models, reflecting reduced headroom, included endocrine (OmicFormer AUC = 0.782, LightGBM = 0.771; Δ = +0.011; +4.59% relative improvement; p = 4.09 × 10⁻³; n = 39) and Ophth & ENT (OmicFormer = 0.766, LightGBM = 0.747; Δ = +0.019; +7.49%; p = 3.71 × 10⁻²; n = 10), where absolute improvements were more modest and statistical support varied by chapter. Detailed per-chapter and per-baseline comparisons are provided in the Supplementary Figure 1 and 2.

### OmicFormer enables accurate prediction on health-related phenotypes

In addition to binary disease prediction, we evaluated OmicFormer on health-related phenotype prediction for 900 quantitative traits, using five baselines: LightGBM, TabMap, ridge regression, 1D-CNN, and TabNet. Model selection was based on Pearson correlation on the validation set, and we reported test-set correlation as the primary metric (Fig. 3a). Detailed trait-level results for proteomics, metabolomics, and brain IDPs are provided in Supplementary Table 4-6. OmicFormer achieved highly significant average improvements across all modalities, and trait-level analysis further revealed that these gains translated into substantial and consistent improvements for a wide range of individual quantitative traits (Fig. 3b).

**Figure 3.**
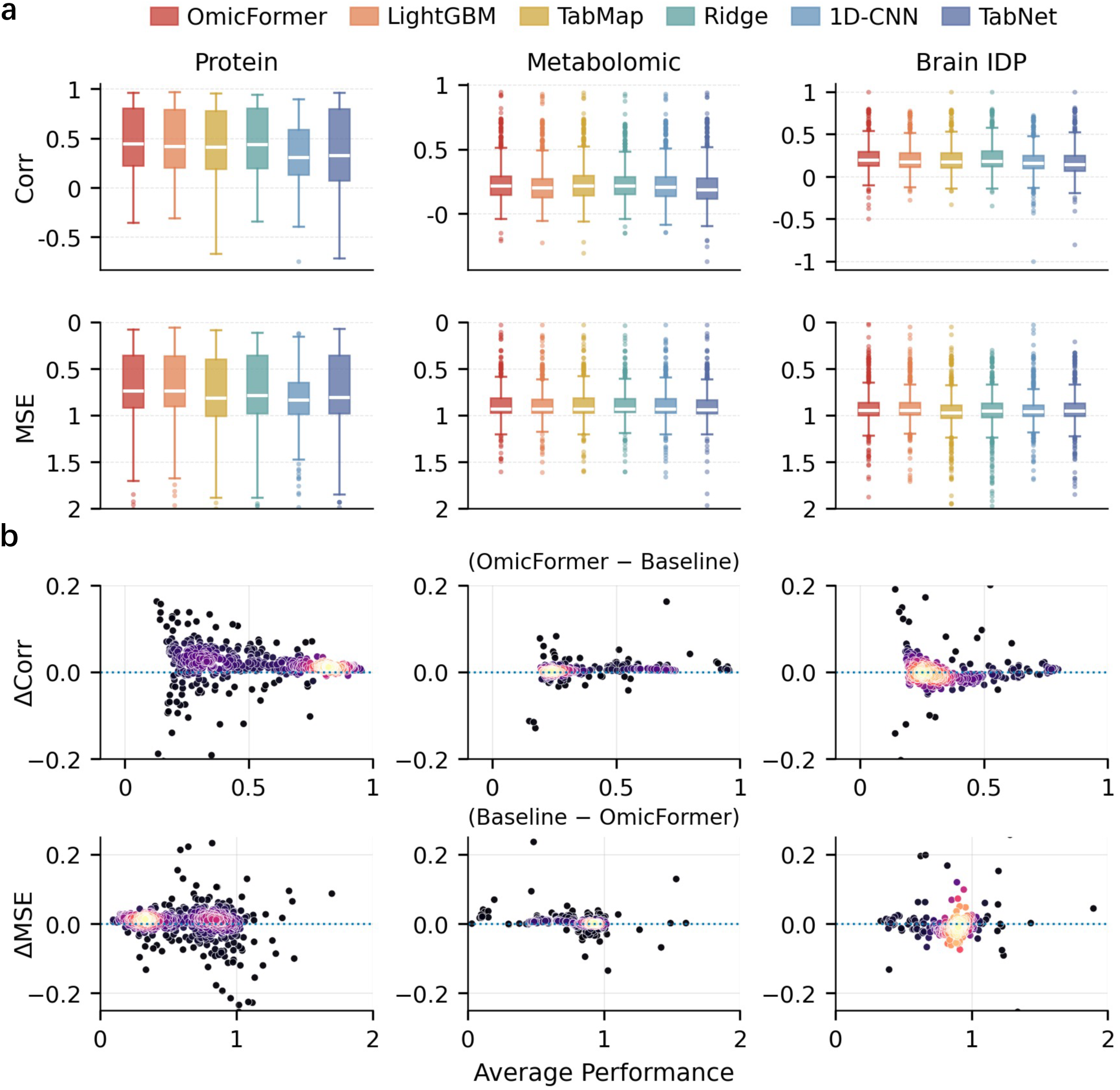
Continuous trait prediction performance and improvement of OmicFormer across omics datasets. a. Predictive performance across modalities. Boxplots display Pearson correlation (top) and mean squared error (MSE, bottom; inverted axis for visual alignment) for proteomics, metabolomics, and imaging-derived phenotypes (IDPs). OmicFormer consistently outperforms all baseline methods (LightGBM, TabMap, TabNet, Ridge regression, and 1D-CNN), achieving higher mean correlation, lower MSE, and reduced variance across traits. b. Per-trait relative improvement over the strongest baseline for each modality. Differences Δ = OmicFormer − baseline are shown for correlation (top) and MSE (bottom), where positive values indicate improvement.

In proteomics, OmicFormer achieved the highest mean correlation among the compared methods, reaching r = 0.486. This significantly outperformed the strongest baseline, ridge regression (r = 0.469), corresponding to a 3.60% relative improvement (p = 2.03 × 10⁻²⁸). OmicFormer also showed clear advantages over other competitive baselines, including LightGBM (r = 0.468; +3.96% relative, p = 3.31 × 10⁻⁵³), TabMap (r = 0.460; p = 1.27 × 10⁻⁷⁷), and TabNet (r = 0.412; p = 8.08 × 10⁻⁸⁹), indicating broad and consistent improvements. At the individual trait level, these highly significant average gains were demonstrated by pronounced improvements for numerous traits. Representative gains included speed of sound through acetate (0.320 to 0.590), cholesterol (0.119 to 0.252), and vertical cup to disc ratio (0.156 to 0.313).

In metabolomics, OmicFormer again achieved the highest mean correlation, with r = 0.265. Compared with the strongest baseline, TabMap (r = 0.260), this corresponded to a 1.85% relative improvement (p = 1.93 × 10⁻¹³). OmicFormer also exceeded LightGBM (r = 0.248; +6.85% relative, p = 3.70 × 10⁻⁵¹), 1D-CNN (r = 0.252; p = 2.36 × 10⁻⁴⁴), and ridge regression (r = 0.253; p = 1.50 × 10⁻²⁴), demonstrating consistent gains in quantitative trait prediction from metabolomic profiles. At the trait level, notable improvements were seen for acetate (0.622 to 0.784), microalbumin in urine (0.188 to 0.224), and vitamin B6 (0.119 to 0.140).

In brain IDPs, OmicFormer attained a mean correlation of r = 0.230, ranking first among all evaluated methods. This is the same as the strongest competing baseline, ridge regression (r = 0.230), while showing substantially better performance than LightGBM (r = 0.215; +7.13% relative, p = 9.63 × 10⁻⁴²) and TabMap (r = 0.213; p = 1.48 × 10⁻⁴⁶). At the trait level, representative gains included heel bone mineral density T-score (0.113 to 0.235), retinal pigment epithelium thickness at central subfield (right) (0.065 to 0.288), and saturated fatty acids (0.108 to 0.180).

### Stratified performance of OmicFormer across phenotype categories

When stratifying continuous outcomes by phenotype categories, OmicFormer’s gains follow a clearer, hierarchical pattern. Using proteomics prediction as an example, for blood assays, the mean correlation increased from r=0.668 (LightGBM) to r=0.683 (OmicFormer), corresponding to an absolute gain of +0.015 (+2.30% relative; p = 3.10 × 10⁻⁷). For NMR metabolomics, performance improved from r=0.757 to r=0.768 (Δr = +0.011; +1.41%; p = 1.50 × 10⁻³²), indicating that even in high signal-to-noise regimes, OmicFormer provides statistically reliable additional benefits.

Notably, OmicFormer also yields substantial improvements in phenotype domains tied to lifestyle and subjective measures. Diet by 24-hour recall shows a marked uplift from r=0.184 to r=0.228 (Δr = +0.044; +23.80%; p = 1.99 × 10⁻⁸), while physical activity measurement increased from r=0.325 to r=0.355 (Δr = +0.031; +9.41%; p = 1.08 × 10⁻¹⁰).

Across several additional categories, OmicFormer similarly outperforms baselines. For example, physical measures improve from r=0.564 (LightGBM) to r=0.581 (OmicFormer) (Δr = +0.017; +2.98%; p = 6.38 × 10⁻⁷), and local environment improved from r=0.146 to r=0.173 (Δr = +0.027; +18.65%; p = 1.20 × 10^-3^). In contrast, for a small number of categories including eye measures and work environment, absolute correlations remain low across methods and gains are less uniform, reflecting persistent challenges in phenotypes that are noisier, sparsely encoded, or measured with coarser instruments. Detailed category-wise comparisons and paired significance summaries are provided in the Supplementary Figure 3 and 4.

**Figure 4.**
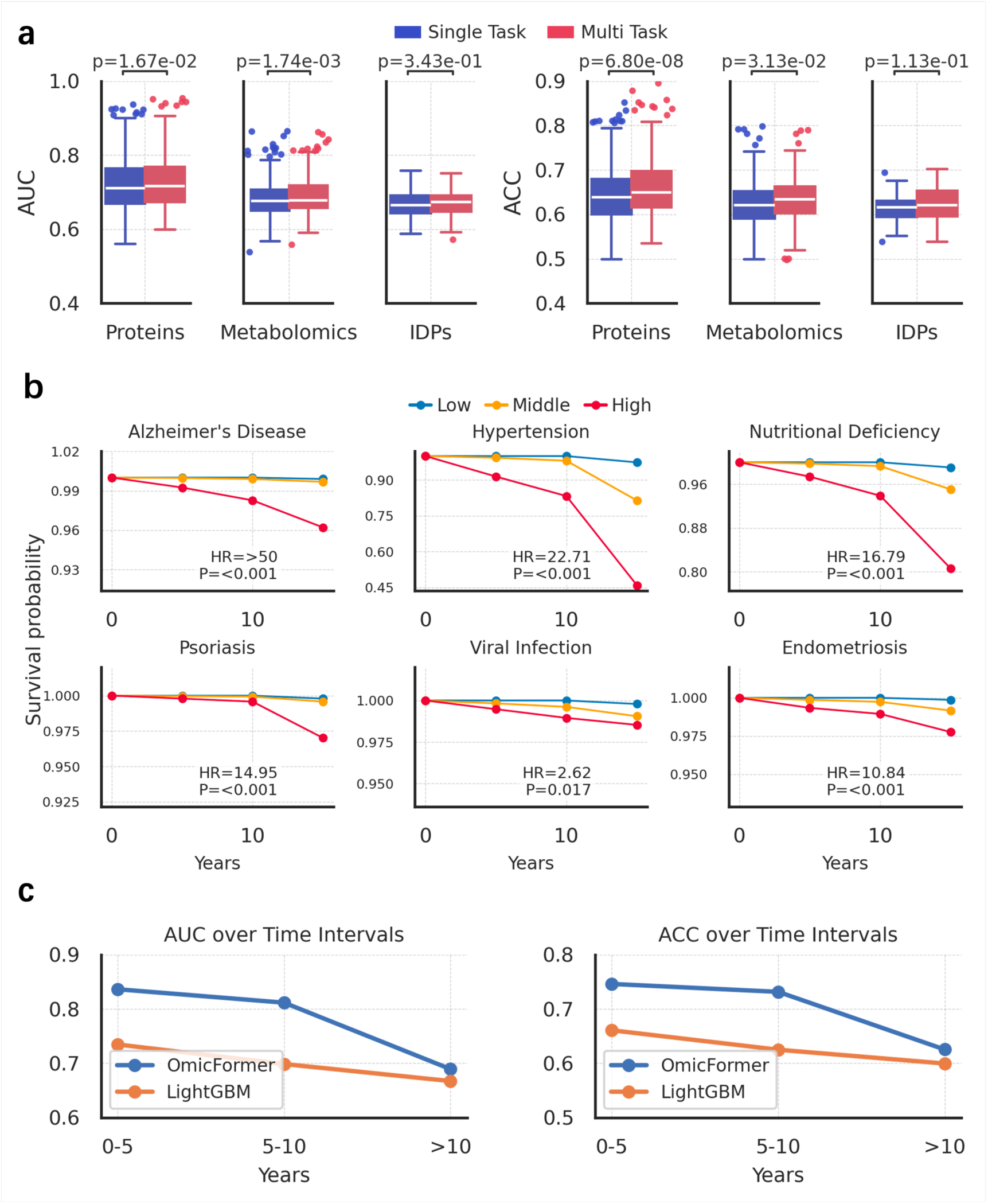
Multi-task learning benefits, risk stratification, and temporal stability of OmicFormer. a. Single-task versus multi-task performance across modalities. Boxplots show AUC (left) and ACC (right) per disease. Multi-task learning consistently improves performance in proteomics and metabolomics, with more moderate gains observed in imaging-derived phenotypes (IDPs). b. Kaplan–Meier risk stratification. Survival curves are stratified by predicted risk groups (low, middle, and high) based on OmicFormer-derived probabilities for representative diseases, demonstrating clinically meaningful separation between risk strata. c. Temporal robustness of OmicFormer prediction across follow-up horizons. AUC performance is compared between OmicFormer and LightGBM across three intervals (0–5, 5–10, and >10 years).

### Multi-task learning further enhances OmicFormer

We next evaluated whether multi-task learning could further improve OmicFormer by explicitly modeling statistical dependencies among prediction targets and leveraging physiological comorbidity structure. This design encourages representation sharing and improved generalization. For diseases, we estimated a pairwise comorbidity probability matrix from the training set and, for each target disease, selected the fifty most comorbid conditions as auxiliary tasks (Methods). All tasks shared a common Transformer encoder, with task-specific classification heads for prediction. The resulting performance comparisons are summarized in Fig. 4a, and further data and cohort details are provided in Supplementary Table 7.

In proteomics, multi-task OmicFormer achieved a relative AUC improvement of 1.64% over the single-task counterpart (p = 1.67 × 10⁻²). The gain in accuracy was more pronounced, with ACC improving by 5.43% (p = 6.80 × 10⁻⁸). At the disease level, representative AUC gains included diabetic neuropathy (0.911 to 0.954; +47.81%), vascular dementia (0.647 to 0.777; +36.76%), and hypertensive renal disease (0.924 to 0.951; +35.74%).

In metabolomics, multi-task OmicFormer yielded the largest relative AUC improvement among the three modalities, reaching 3.44% compared with single-task training (p = 1.74 × 10⁻³). The ACC also improved by 3.17% (p = 3.13 × 10⁻²). At the disease level, representative AUC gains included hemiplegia (0.538 to 0.658; +25.93%), cerebral palsy and other paralytic syndromes (0.568 to 0.655; +20.12%), and duodenal ulcer (0.667 to 0.733; +19.72%).

In brain IDPs, multi-task OmicFormer achieved a relative AUC improvement of 1.27% over the single-task model (p = 3.40 × 10⁻¹). ACC increased by 2.84% (p = 1.1 × 10⁻¹). Although the direction of improvement remained positive, these gains were more modest and did not reach statistical significance, suggesting that the benefit of multi-task learning in brain imaging IDPs may be weaker than that observed in proteomics and metabolomics. At the disease level, representative AUC gains included type 2 diabetes (0.691 to 0.745; +17.54%), disorders of gallbladder, biliary tract and pancreas (0.589 to 0.651; +15.05%), and acute renal failure (0.642 to 0.693; +14.40%).

These results indicate that multi-task learning provides a consistent benefit for OmicFormer, with the clearest gains observed in proteomics and metabolomics, where modeling inter-disease dependencies enhanced prediction across numerous individual conditions. Notably, this multi-task formulation is naturally supported by shared-representation neural architectures such as OmicFormer, whereas extending tree-based models to the same setting is less straightforward, highlighting a practical advantage of the proposed framework.

### Risk stratification and survival analysis

To evaluate the clinical interpretability of OmicFormer-derived risk scores, we performed Kaplan–Meier survival analysis^26^ using predicted probabilities from the proteomics-based disease prediction. Test-set participants were stratified into low-, intermediate-, and high-risk groups according to predicted risk (Methods), and disease-free survival was compared over a fixed follow-up window (Fig. 4b).

For many diseases, the high-risk group consistently exhibited lower disease-free survival than the intermediate-and low-risk groups, with the separation between groups generally widening over time. The clearest separation was observed for outcomes such as essential hypertension (HR = 22.71, p < 0.001), polymyalgia rheumatica (HR = 32.09, p < 0.001), nutritional deficiency (HR = 16.79, p < 0.001), psoriasis (HR = 14.95, p < 0.001), and postoperative endocrine disorders (HR = 10.84, p < 0.001) (Fig. 4c). More moderate but still significant separation was observed for fistula (HR = 2.35, p = 0.038) and viral infection–related outcomes (HR = 2.62, p = 0.017). Overall, these results indicate that OmicFormer-derived risk scores provide clinically meaningful stratification of future disease risk.

### Future disease prediction performance across follow-up intervals

We evaluated temporal robustness of OmicFormer using proteomics-based future disease risk prediction as a representative example. Across all models, predictive performance declined with increasing follow-up duration.

OmicFormer achieved average AUCs of 0.836, 0.812, and 0.689 for prediction horizons of <5 years, 5–10 years, and >10 years, respectively, with corresponding ACCs of 0.746, 0.732, and 0.625. This progressive attenuation likely reflects increased uncertainty, cumulative noise, and competing risks inherent in long-term event prediction.

Despite the overall performance decay, OmicFormer consistently outperformed LightGBM across all follow-up intervals. Under the same evaluation framework, LightGBM achieved AUCs of 0.735, 0.699, and 0.667, and ACCs of 0.661, 0.625, and 0.600 (Fig. 4c).

### Biologically interpretable features contribute to OmicFormer predictions

We evaluated the interpretability of OmicFormer using proteomics-based future disease prediction as a representative case study. Feature importance was quantified using grad-cam, a widely adopted attribution method for deep neural networks^27^. A protein was counted as a single contribution if it ranked among the top five for a given disease. Fig. 5a summarizes the cumulative frequency of these top-five proteins across all diseases.

**Figure 5.**
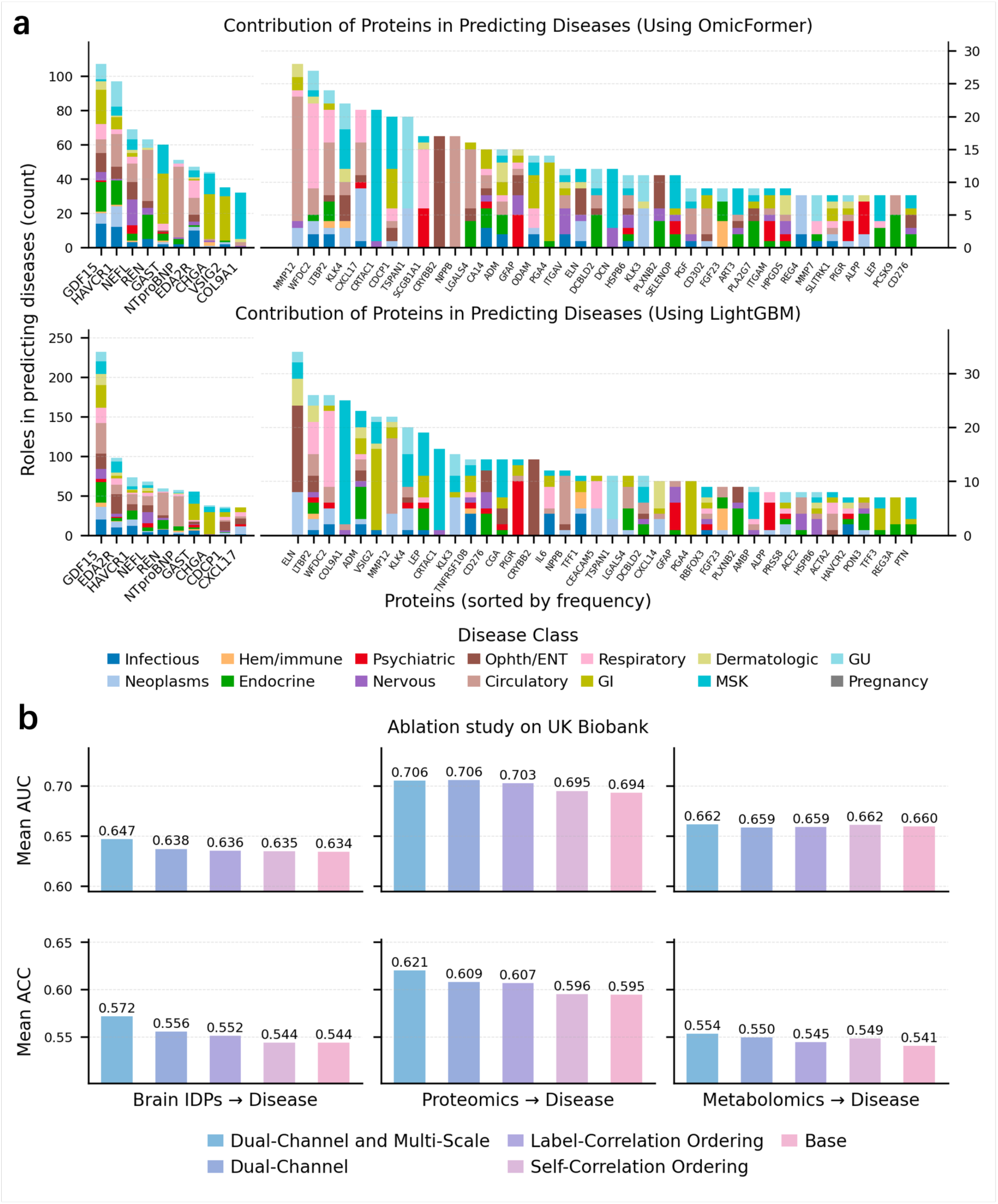
Feature contribution patterns and ablation analysis of OmicFormer. a.Feature contribution across proteomics disease prediction tasks. Bar plots show per-protein contribution frequency across 450 diseases for OmicFormer (top) and LightGBM (bottom), revealing that OmicFormer identifies a broader spectrum of biologically relevant biomarkers while preserving core signals captured by tree-based methods. b.Ablation study on OmicFormer architectural design. Mean AUC (top) and ACC (bottom) are shown across modalities under different configurations. The full model incorporating both dual-channel priors and multi-scale patch embedding achieves the best performance, followed by partial variants, while the base model without structural priors performs worst, confirming the complementary contributions of each component.

Several proteins, most prominently GDF15, HAVCR1, and NT-proBNP, were consistently prioritized across multiple disease categories, forming a set of high-frequency, broadly shared biomarkers. These findings are concordant with established biological knowledge: GDF15 is linked to systemic inflammation and metabolic dysregulation^28^; HAVCR1 reflects kidney injury and immune activation^29^; and NT-proBNP is a canonical marker of cardiovascular risk^30^. The alignment with prior clinical and epidemiological evidence supports the biological plausibility of the representations learned by OmicFormer.

For comparison, Fig. 5a also presents protein contribution distributions derived from LightGBM feature importance score. Tree-based models tended to concentrate importance on a small set of highly frequent, linearly separable features. In contrast, OmicFormer retained these core proteins while additionally identifying a broader spectrum of moderate-frequency, disease-specific markers. This resulted in a more structured importance landscape in which shared and condition-specific biomarkers coexisted.

The grad-cam framework also extends to brain imaging IDPs. In this modality, each IDP was counted once for a given disease if it ranked among the top five contributors. Repeated selection of regional brain volumes, cortical thickness measures, and white-matter integrity indices across neurological and systemic diseases revealed imaging markers associated with brain–body axis risk. Comprehensive cross-modality biomarker summaries are provided in Supplementary Figure 5.

### Ablation study on OmicFormer design

To evaluate the contribution of the dual-channel correlation priors and the multi-scale patch embedding to disease prediction, we conducted a comprehensive ablation study across three omic modalities (Fig. 5b).

Using the full model (Dual-Channel and Multi-Scale) as the reference, we first examined the effect of single-channel feature ordering. Retaining only the label-correlation ordering or only the self-correlation ordering led to consistently lower average AUCs across all three modalities. For example, in the proteomics tasks, the single-channel variants achieved mean AUCs of 0.687 (retaining only the label-correlation ordering) and 0.674 (retaining only the self-correlation ordering), compared with 0.697 for the full model. Similar trends were observed for brain IDPs and metabolomics, indicating that the two correlation priors provide complementary information.

Next, we evaluated a Dual-Channel (no Multi-Scale) variant that preserves both ordering channels but replaces the multi-scale patch embedding with a single-scale projection. This modification resulted in a consistent performance drop of approximately 0.005–0.01 mean AUC across modalities, with more pronounced degradation in the proteomics and metabolomics tasks. These results suggest that multi-scale modeling is critical for capturing dependencies at different receptive-field ranges in high-dimensional tabular features.

Finally, compared with a baseline model without any feature reordering, most correlation-based variants achieved superior performance, confirming that correlation-driven one-dimensional feature reordering itself constitutes an effective, parameter-free structural prior. Overall, label-correlation ordering and self-correlation ordering reshape the feature space from task-relevance and structural-dependence perspectives, respectively, while multi-scale patch embedding further enhances local semantic aggregation. The synergy of these components accounts for the observed performance gains across multimodal disease prediction tasks.

### External generalizability of OmicFormer on GNPC

We conducted external validation by training disease prediction models on UKB proteomics and evaluating performance on GNPC^24^ (Fig. 6, Supplementary Figure 6, Supplementary Table 8).

**Figure 6.**
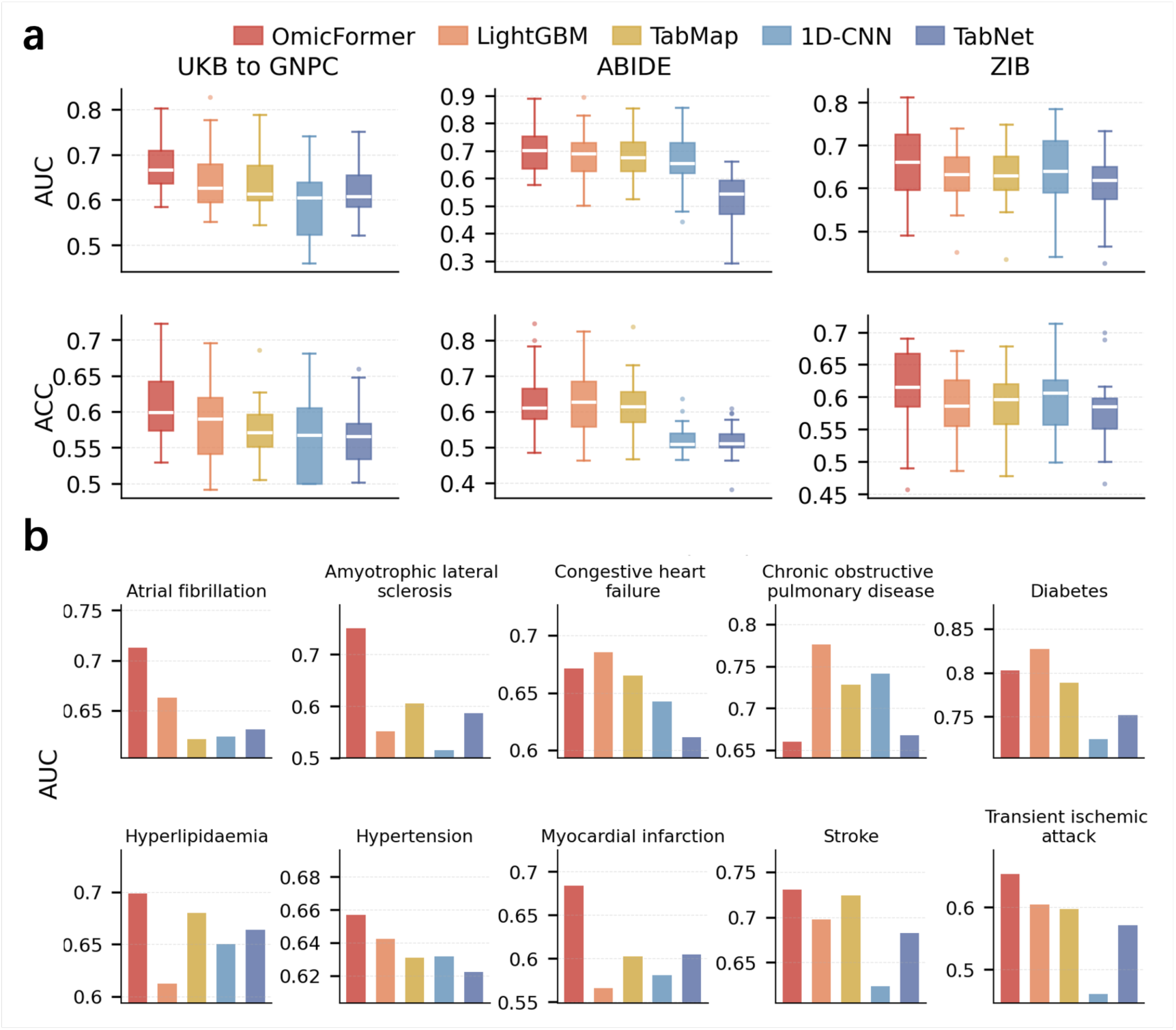
External validation and cross-cohort generalization of OmicFormer. a. Cross-cohort and leave-one-site evaluation. Boxplots show AUC (top) and ACC (bottom) under external validation settings. Left: models trained on UK Biobank and tested on the independent GNPC proteomics cohort. Middle and right: leave-one-site-out evaluation on ABIDE (autism) and ZIB (schizophrenia) neuroimaging datasets, respectively. b. Disease-specific external validation on GNPC. AUC performance is shown for ten representative diseases for which at least one method achieved AUC > 0.65, comparing OmicFormer against the 4 baselines for each disease using UK Biobank-trained models tested on GNPC.

In the UKB-trained and GNPC-tested proteomics prediction, OmicFormer achieved a mean AUC of 0.675, corresponding to an 8.30% relative improvement over LightGBM, the strongest baseline on average (AUC=0.646), and a mean ACC of 0.613, representing a 6.73% relative improvement over LightGBM (ACC=0.585). Consistent with the boxplot distributions (Fig. 6a), OmicFormer showed a higher central tendency across diseases with comparable variability, suggesting a broadly distributed advantage under external cohort shift.

Fig. 6b further presents representative disease-level results for clinically meaningful prediction tasks, defined as diseases for which at least one compared model achieved AUC > 0.65 on GNPC. For each disease, OmicFormer was compared against the strongest baseline for that specific task. Relative to the disease-specific best baseline, OmicFormer showed the largest gain for amyotrophic lateral sclerosis (ALS), improving AUC from 0.605 to 0.750 (Δ = 0.145, +36.71%, p < 2.00 × 10^-4^). Significant gains were also observed for myocardial infarction, improving from 0.605 to 0.684 (Δ = 0.079, +20.00%, p < 2.00 × 10^-4^), hypertension, from 0.639 to 0.657 (Δ = 0.018, +4.99%, p = 3.98 × 10^-4^), and hyperlipidaemia, from 0.680 to 0.698 (Δ = 0.018, +5.63%, p = 4.96 × 10^-2^). For several additional diseases, including transient ischaemic attack, atrial fibrillation, congestive heart failure, and stroke, OmicFormer also achieved slightly better performance than the strongest baseline. OmicFormer achieved a similar performance with strongest baseline for diabetes or chronic obstructive pulmonary disease.

### External generalizability of OmicFormer on brain imaging-derived phenotypes

We further assessed independent-site generalization on ABIDE (ASD)^20^ and ZIB (schizophrenia)^25^ using a leave-one-site-out (LOSO) protocol (Fig. 6, Supplementary Table 9-10).

In the ABIDE autism prediction, OmicFormer achieved a mean AUC of 0.699, compared with 0.681 for LightGBM, corresponding to a 5.55% relative improvement (p = 2.02 × 10⁻²). Site-level analysis showed AUC improvements at several relatively large held-out sites, including ABIDEII-GU_1 (n = 99, +0.112), NYU (n = 171, +0.040), UM_1 (n = 82, +0.039), USM (n = 61, +0.032), and UCLA_1 (n = 55, +0.054). For accuracy, OmicFormer reached a mean ACC of 0.631, compared with 0.616 for LightGBM, corresponding to a 4.09% relative improvement. Accuracy gains were likewise observed at several higher-sample sites, including ABIDEII-GU_1 (n = 99, +0.140), NYU (n = 171, +0.027), UM_1 (n = 82, +0.062), USM (n = 61, +0.014), and UCLA_1 (n = 55, +0.017). Across the remaining sites, performance was broadly comparable, with mixed but generally modest differences.

In the ZIB schizophrenia prediction, OmicFormer achieved a mean AUC of 0.672, compared with 0.637 for LightGBM, corresponding to a 9.43% relative improvement. Site-level analysis showed statistically significant AUC improvements at three sites: COBRE (n=165, +0.094, 29.74% relative improvement, p = 1.48 × 10^-2^), MCIC (n=203, +0.135, 35.14% relative improvement, p = 2.40 × 10^-3^), and NUSDAST (n=250, +0.143, 33.93% relative improvement, p = 6.00 × 10^-4^). At the remaining sites, performance was broadly comparable, with no statistically significant differences. For accuracy, OmicFormer reached a mean ACC of 0.615, compared with 0.592 for LightGBM, corresponding to a 5.69% relative improvement.

Collectively, across cross-cohort and multi-site evaluations, OmicFormer consistently exceeded or matched strong baselines. These findings support the robustness and cross-domain generalization capability of the proposed architecture.

## Discussion

The expansion of high-dimensional omics data in biomedical research has created both opportunities and challenges for precision medicine. While traditional tree-based methods remain strong baselines for tabular data, their inherent limitations, e.g., greedy splitting, inability to model global feature interactions, and performance degradation under distribution shifts, constrain their utility across diverse cohorts and multimodal settings. Here, we introduce OmicFormer, a Transformer-based architecture that explicitly encodes statistical structure into the input representation through dual-channel correlation priors, enabling more effective learning of both local and long-range dependencies in omics data.

Across extensive evaluation on the UK Biobank encompassing 450 diseases and 900 quantitative traits, OmicFormer consistently outperforms strong baselines. Given the large number of phenotypes evaluated, spanning hundreds of heterogeneous tasks across three omics modalities, achieving statistically significant average improvements represents a particularly stringent benchmark. The heterogeneity in prediction difficulty, underlying biological mechanisms, and signal-to-noise ratios across such a diverse set of phenotypes means that even modest average gains are nontrivial, as any single method is unlikely to uniformly excel across all tasks. In this context, the consistent and highly significant average improvements delivered by OmicFormer, together with its widespread gains at the individual phenotype level, underscore the robustness and generalizability of the proposed approach across a broad spectrum of health-related prediction tasks.

Another critical finding is OmicFormer’s robust external generalization under distribution shift. In proteomics-based validation on the independent GNPC cohort, OmicFormer maintained superior performance compared with tree-based methods, which exhibited substantial degradation. Similarly, in neuroimaging-based leave-one-site-out validation on ABIDE and ZIB, OmicFormer demonstrated both higher mean performance and reduced cross-site variability.

A central insight of our model design is that the ordering of features in the input sequence matters fundamentally for tabular transformer models. The attention mechanisms benefit when semantically related features are positioned nearby due to the use of positional encoding. By reordering features according to their correlation with the target label, OmicFormer creates a task-relevant topology that allows local receptive fields to capture coherent predictive signals. Additionally, the complementary feature–feature correlation channel, optimized via optimal transport, encodes intrinsic biological dependencies, such as protein interaction modules, metabolomic pathway relationships, or brain structural connectomes, directly into the sequence layout. Therefore, this dual-channel design transforms a generic tabular input into a semantically structured representation without increasing computational complexity or requiring external knowledge graphs.

The integration of multi-scale patch embedding further enhances representation learning by capturing patterns at varying receptive fields. This design, inspired by hierarchical vision transformers^31–34^, proves particularly valuable for omics data where predictive signals manifest at multiple scales: local motifs (e.g., local protein networks), medium-range dependencies (e.g., pathway-level coordination), and global feature interactions. Our ablation studies confirm that both correlation channels contribute complementary information and that multi-scale modeling provides consistent gains across modalities, with the full model achieving best performance improvements.

Several limitations and future directions warrant discussion. First, while correlation-based ordering provides a strong structural prior, it assumes stationary feature relationships that may vary across biological contexts or disease states. Developing adaptive ordering mechanisms that condition on task or cohort characteristics could further improve flexibility. Second, the current framework treats each modality independently; extending OmicFormer to explicitly model cross-modal interactions, for example, between proteomics and imaging phenotypes, represents a natural next step toward integrated multi-omics prediction. Third, while Grad-cam attribution offers model interpretability, more rigorous causal inference frameworks could help distinguish correlation from causation in identified biomarkers.

In conclusion, OmicFormer is a principled approach to omics-based prediction by embedding dual-channel correlation priors into a Transformer architecture. Through systematic evaluation across proteomics, metabolomics, and neuroimaging modalities, we demonstrate consistent performance improvements, robust cross-cohort generalization, and biologically interpretable representations. By showing that explicit statistical structure can enhance deep learning for tabular omics data, this work provides both a practical tool for precision medicine and a conceptual framework for integrating domain knowledge into representation learning.

## Data availability

The individual-level data used in the present study were obtained from UK Biobank (https://www.ukbiobank.ac.uk/), the Global Neurodegeneration Proteomics Consortium (GNPC; https://www.neuroproteome.org/), the Autism Brain Imaging Data Exchange (ABIDE; https://fcon_1000.projects.nitrc.org/indi/abide/), and the Zhangjiang International Brain BioBank (ZIB; https://zib.fudan.edu.cn/). Access to these datasets is subject to the respective data access and approval procedures of each resource.

## Code availability

The code to reproduce the results can be accessed at https://github.com/PB21000283/OmicFormer.git.

## Author contributions

W.G, W,C, H,J designed the project, H,J, C.Y collected and analysed the data, wrote the manuscript. All authors discussed the results and reviewed the manuscript.

## Acknowledgements

This work was supported by Noncommunicable Chronic Diseases-National Science and Technology Major Project (2025ZD0546300) and the National Natural Science Foundation of China (Grant No. 82402379).The UKB analyses were conducted using the UKB Resource under application no. 19542.

## Methods

### Overall architecture of OmicFormer

OmicFormer is a correlation-prior–guided dual-channel one-dimensional Transformer framework for unified modeling of multi-omic tabular data, including proteomics, metabolomics, and brain IDPs. The core design principle is to incorporate explicit statistical structure priors into both sequence construction and representation learning while preserving the native feature-axis semantics of tabular inputs.

Specifically, OmicFormer constructs two complementary input channels. The first channel adopts a label-guided ordering strategy, where features are ranked by the absolute value of their Pearson correlation coefficient with the target variable, encouraging highly informative features to form locally concentrated regions along the one-dimensional sequence. The second channel captures intrinsic feature dependencies by leveraging the feature–feature correlation matrix using optimal transport and performing a one-dimensional projection that promotes locality among strongly correlated features, thereby making statistical interactions more explicit in the sequence geometry.

Each channel is processed through a multi-scale 1D convolutional patch embedding module, which extracts representations across receptive fields ranging from local to global scales. The resulting patch tokens are fed into a shared Transformer encoder, where self-attention models long-range interactions among features. To integrate complementary information across channels, we introduce a learnable gating mechanism that adaptively assigns channel-wise weights prior to encoding. The fused representation is subsequently passed to a task-specific prediction head to generate the final output.

Overall, the architecture is organized around three core components: (1) The channel one with label-guided feature ordering; (2) The channel two with correlation-guided feature organization; (3) The multi-scale patch embedding combined with Transformer encoding. Together, these components establish a unified, correlation-aware pipeline that enables structured and globally informed dependency modeling in omics tabular data.

### Auxiliary task selection for multi-task learning

To mitigate negative transfer in multi-task training, OmicFormer adopts relevance-based auxiliary task selection rather than indiscriminately training on all available tasks. For binary disease prediction, we quantify comorbidity-based relatedness between a target disease *t* and a candidate disease *k*. Let *n*_t_, *n*_k_, and *n*_t,k,_ denote the numbers of positive cases for disease *t*, disease *k*, and both jointly. We define a symmetric comorbidity score:

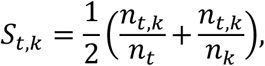

which measures normalized co-occurrence relative to each disease’s prevalence. Candidate diseases are ranked by *S*_t,k,_; those with *S*_t,k_ > 0.2 are retained, with at most 50 auxiliary tasks selected. For continuous phenotype prediction, we compute Pearson correlations *r*_t,k_ = corr(*y*_t_, *y*_k_) between phenotype labels and rank candidates by ∣ *r*_t,k_ ∣. Tasks satisfying ∣ *r*_t,k_ ∣> 0.2 are retained, again capped at 50 auxiliary tasks. This relevance-driven selection prioritizes statistically coupled outcomes while suppressing weakly related tasks, thereby improving stability and reducing negative transfer in multi-task learning.

### Feature ordering and dual-channel inputs

Let *X* ∈ ℝ*^N^*^×p^ denote the tabular omics matrix with *N* subjects and *p* features, where *x*_i,j(_ is the value of feature *j*for subject *i*. The target label is *y* ∈ ℝ*^N^*(binary disease status or a continuous trait). OmicFormer constructs two complementary 1D feature sequences (channels) to inject statistical priors into the input representation.

### Channel 1: Label-guided ordering (feature–label correlation prior)

For each feature *j* ∈ {1,…,*p*}, we compute the Pearson correlation between feature *X*_:j(_and label *y*:

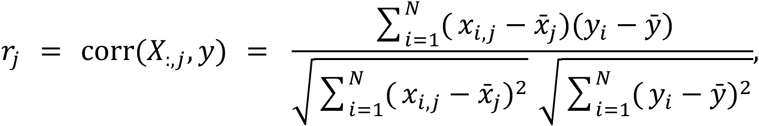

where 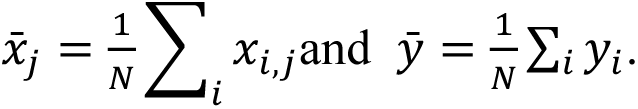

We then sort features by decreasing absolute correlation magnitude:

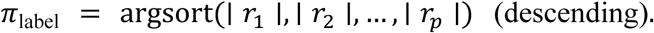

This induces a reordered sequence

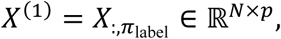

which concentrates label-informative features early in the 1D order, enabling convolutional or local attention blocks to more easily capture predictive patterns.

### Channel 2: Correlation-guided feature reorganization using Gromov–Wasserstein optimal transport^12^

Channel 2 aims to preserve feature–feature dependency by reordering features so that mutually correlated features become neighbors in the 1D sequence.

### **(1)** Feature–feature correlation and distance matrix

We compute the Pearson correlation matrix *R* ∈ ℝ^p×p^:

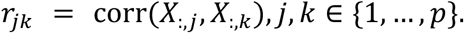

We convert correlation to a correlation-distance matrix *C* ∈ ℝ^p×p^using:

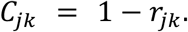

### **(2)** 1D grid distance matrix

Define a 1D grid with *m*positions:

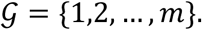

The grid distance matrix 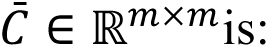

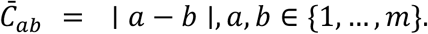

This encodes the geometry of a 1D line, i.e., neighbors in index have small distance.

### **(3)** Gromov–Wasserstein optimal transport alignment

We aim to align the metric structure of the feature space (*C*)and the 1D grid space 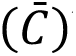 by solving a Gromov–Wasserstein (GW) optimal transport problem.

Let *u* ∈ Δ_m_ and *v* ∈ Δ_p_ be marginal distributions over grid positions and features. In our construction, we use uniform distributions:

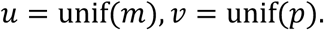

We solve for a coupling matrix 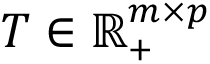 satisfying:

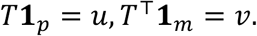

The GW objective minimizes the discrepancy between pairwise distances in the two spaces:

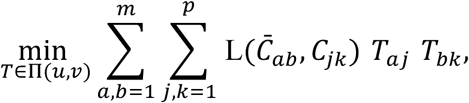

where Π(*u*, *v*) = {*T* ≥ 0 ∣ *T***1** = *u*, *T*^4^**1** = *v*}and *L*(⋅,⋅)is a loss comparing distances.

Square loss (common GW form):

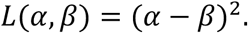

In practice, the solver normalizes distance matrices (as in your code C /= C.mean()), initializes *T*: = *uv*^4^, and optimizes *T*using either:

Exact OT (emd) when *ε* = 0:

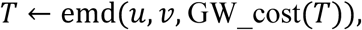

Entropic-regularized Sinkhorn when *ε* > 0:

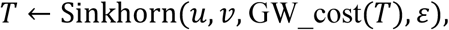

iterating until convergence ∥*T* − *T*_prev_ ∥;≤ tol.

### **(4)** From soft coupling to permutation (Hungarian assignment)

The GW solver yields a soft coupling *T*. To obtain a one-to-one feature ordering (permutation), we apply a linear sum assignment (Hungarian algorithm):

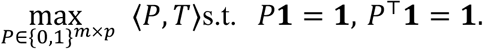

This produces a permutation-like binary matrix *P*(your code linear_sum_assignment(-T_prime)), which defines the final reordered feature sequence:

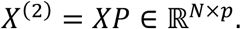

Thus, correlated features (small *C*_jk_) are mapped to nearby grid indices (small ∣ *a* − *b* ∣), yielding a structure-friendly 1D layout.

### **(5)** Dual-channel fusion with learnable gating (brief transition sentence)

Finally, OmicFormer feeds both sequences, *X*^(1)^and *X*^(2)^, into a dual-branch encoder and applies a learnable gating mechanism to adaptively fuse the two priors before multi-scale 1D ViT encoding. Specifically, the fused representation is defined as

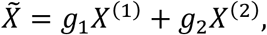

where the channel weights *g*_1_ and *g*_2_ are obtained from a softmax-normalized learnable gate,

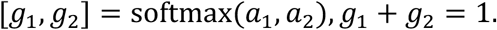

This mechanism allows the model to dynamically balance the relative contributions of the two priors, thereby emphasizing more informative signals while reducing the influence of noisier or less relevant ones. The fused representation 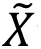 is then passed to the multi-scale 1D ViT backbone to capture local-to-global dependencies.

### Multi-scale patch embedding with token alignment

Let the reordered 1D input sequence be *X* ∈ ℝ^B×C×L^, where *B* denotes the batch size, *C* the number of input channels, and *L* the feature length. To capture dependencies across multiple spatial scales, we employ *S*parallel 1D convolutional branches with kernel sizes 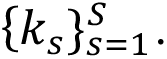. Each branch computes

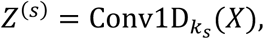

with elements

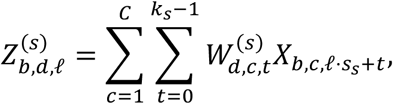

where *s*_s_ = max (1, *k*_s_/2) is the stride. Larger kernels correspond to broader receptive fields but typically yield shorter token sequences *L*_S_. To enable cross-scale fusion, each branch is resampled to a common length *L*_max_ = max _S_ *L*_S_ via 1D linear interpolation,

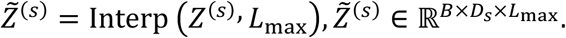

The aligned features are concatenated along the embedding dimension,

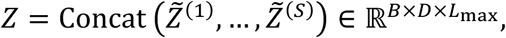

and reshaped into token embeddings *X*_tokens_ ∈ ℝ^B×N×D^, which serve as the Transformer input. This multi-scale formulation integrates fine-grained and global contextual information while preserving a consistent token resolution across branches.

### Transformer encoding with positional priors

The token sequence is processed by a stack of multi-head self-attention and feed-forward layers to model long-range interactions. Given *H*^(0)^ = *X*_tokens_, each Transformer layer computes

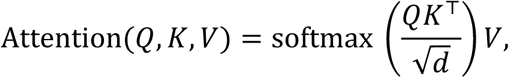

followed by position-wise feed-forward transformations. To encode structural priors derived from correlation-guided feature reordering, we incorporate 1D sinusoidal positional embeddings defined as

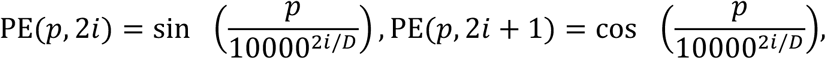

where *p*denotes the token index under the correlation-informed permutation. The encoder input becomes

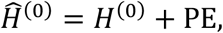

thereby embedding statistical ordering priors directly into the positional geometry of the sequence. This integration of correlation-aware ordering and sinusoidal encoding introduces an inductive bias that aligns attention mechanisms with the intrinsic dependency structure of the data.

### Prediction heads and training objectives

OmicFormer adopts task-specific pooling strategies to accommodate classification and regression objectives. For classification, a learnable [CLS] token is prepended to the token sequence before Transformer encoding. Let *H*^(L)^ ∈ ℝ^B×(N+1)×D^ denote the final-layer representations; the [CLS]embedding 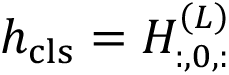 is passed to an MLP classifier to produce logits 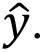 For continuous prediction, instead of relying on a single token, we aggregate global information by flattening all token embeddings,and feeding the concatenated vector into a fully connected MLP regressor. For classification, we minimize a class-weighted cross-entropy loss to address imbalance,

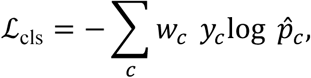

where *w*_C_ is the inverse-frequency weight for class *c*. For regression, we use mean squared error,

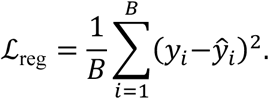

In the multi-task setting, the total objective is a weighted sum over the primary task and selected auxiliary tasks,

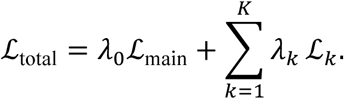

Training is performed using AdamW with cosine annealing warmup restarts. Dropout is applied within the patch embedding and Transformer blocks, and L2 regularization is incorporated via weight decay to mitigate overfitting and enhance generalization.

### Dataset description

#### UK Biobank (UKB)

This study is primarily based on the UKB, a large-scale prospective population cohort comprising approximately 500,000 participants recruited across the United Kingdom^3,24^. UKB integrates extensive phenotypic, genomic, biochemical, proteomic, metabolomic, and neuroimaging data, enabling systematic investigation of complex disease etiology and population-level health determinants. For proteomics, we use data from the UK Biobank Pharma Proteomics Project (UKB-PPP), in which approximately 54,000 plasma samples were profiled using the Olink Explore 3072 platform, quantifying 2,920 circulating proteins across diverse biological pathways. For metabolomics, we analyze blood-based metabolic biomarkers measured at population scale using the Nightingale Health nuclear magnetic resonance platform. For neuroimaging, we use brain IDPs generated from structural MRI, diffusion MRI, and functional MRI pipelines, including volumetric measures, white matter microstructure indices, and functional connectivity features that have been extensively validated in large-scale neuroimaging research.

#### Global Neurodegeneration Proteomics Consortium (GNPC)

For external validation in proteomics, we used data from the Global Neurodegeneration Proteomics Consortium (GNPC)^24^, a large multi-cohort initiative designed to characterize plasma proteomic signatures associated with neurodegenerative and systemic diseases across diverse populations. The GNPC external test set included 7,289 samples. GNPC aggregates Olink-based proteomic measurements from multiple independent studies, encompassing heterogeneous demographic backgrounds, recruitment settings, and clinical phenotypes, thereby providing a realistic benchmark for cross-cohort generalization under distributional shift. We identified 2,137 proteins shared between the UK Biobank and GNPC to ensure direct molecular comparability. Feature selection was performed exclusively on the UKB training set by ranking proteins according to absolute feature–label Pearson correlation and retaining the top 20% most correlated proteins; this identical feature subset was then applied across all methods during GNPC testing to avoid information leakage. Models were trained and validated solely on UKB and externally evaluated on GNPC without fine-tuning. Disease definitions in GNPC were aligned to UKB ICD-10–based outcomes via a one-to-many semantic mapping to ensure diagnostic consistency, covering 19 clinical outcomes. All methods shared identical random seeds, preprocessing, normalization, and evaluation protocols, and summary statistics were reported only for diseases where at least one method achieved AUC ≥ 0.6 on GNPC to ensure stable and directly comparable external-test analyses.

#### Independent multi-site neuroimaging cohorts (ABIDE and ZIB)

To evaluate cross-site generalizability in neuroimaging settings, we include two independent multi-center cohorts. The Autism Brain Imaging Data Exchange (ABIDE)^20^ is an international consortium aggregating resting-state functional MRI data from 34 acquisition sites to study autism spectrum disorder. The cohort includes 1,779 participants and exhibits substantial heterogeneity in scanner types, acquisition protocols, and demographic composition, making it widely used for assessing robustness under site-level distribution shifts. In addition, we use data from the Zhangjiang Imaging Brain Bank (ZIB)^25^, a multi-site neuroimaging cohort collected across 17 sites, comprising structural and functional imaging-derived phenotypes for psychiatric and neurological research. The ZIB cohort includes 2,949 participants and provides site-diverse imaging data suitable for independent-site validation and cross-center reproducibility assessment. Together, ABIDE and ZIB offer complementary multi-site neuroimaging resources for evaluating cross-site reproducibility and model generalizability.

#### Training and Evaluation

We study two phenotype prediction settings: disease classification and continuous phenotype regression. Disease prediction includes both future incidence risk and prior diagnosis status, formulated as binary case–control classification; regression targets quantitative clinical and biochemical traits. For regression, we use a random 0.8/0.1/0.1 split into training/validation/test sets. For classification, to improve the credibility and comparability of evaluation across diseases, we construct fixed validation and test subsets by sampling 75 cases for validation and 75 cases for testing. For each selected case, we retrieve five controls matched by nearest age within the same sex; controls may repeat across different cases because matching is performed independently per case. For classification, we select checkpoints by validation AUC (early stopping) and report AUC together with balanced ACC on the test set to better reflect discrimination under class imbalance. For regression, we select models by validation Pearson correlation and report corr and mean squared error (MSE) on the test set.

To ensure fair and controlled comparisons, OmicFormer is trained with a fixed hyperparameter configuration chosen in preliminary experiments, focusing the comparison on architectural contributions. For deep-learning baselines (1D-CNN, TabMap, and TabNet), we align key architectural and regularization choices with our model (e.g., embedding dimension and dropout), and train each baseline over five candidate learning rates, reporting the checkpoint with the best validation performance. LightGBM is tuned using 20 rounds of random hyperparameter searches, providing a competitive tuning budget for tree-based baselines while isolating the gains attributable to correlation priors and multi-scale Transformer modeling.

For continuous regression, OmicFormer uses a lightweight prediction head: output tokens are flattened and passed to an MLP to produce the final scalar (or trait-wise) prediction.

For metabolomics-based prediction, including both disease prediction and continuous phenotype prediction, the original sample size was substantially larger (approximately 270,000 individuals). To improve computational efficiency while preserving population-scale diversity, we randomly sampled 60,000 individuals for downstream training and evaluation.

To improve statistical stability and comparability across tasks, we report results on a filtered disease subset. For each modality, we first include only diseases with more than 200 cases, leaving 238, 450, and 442 diseases in the brain IDP, proteomics, and metabolomics settings, respectively. We then apply a performance-based filter and retain diseases for which at least one method achieves AUC ≥ 0.65, resulting in final subsets of 69, 293, and 96 diseases, respectively.For continuous phenotype regression, the original numbers of traits were 900, 899, and 642 for the brain IDP, proteomics, and metabolomics settings, respectively. We further retain only traits for which at least one method achieves a test correlation greater than r=0.1, resulting in final subsets of 716, 796, and 461 traits, respectively.

### Risk stratification and survival curve analysis

To assess the clinical interpretability of model-derived risk scores, we further conducted risk stratification followed by Kaplan–Meier survival analysis^26^ using the OmicFormer model’s predicted probabilities. For each disease, individuals with prevalent conditions at baseline were excluded, retaining only incident cases and censored individuals during follow-up.

Participants in the test set were stratified into low-risk (bottom 20%), intermediate-risk (middle 60%), and high-risk (top 20%) groups based on predicted disease probability. Time-to-event was defined as the duration from baseline to first diagnosis or censoring, and survival curves were estimated and compared within a 15-year follow-up window.

#### Prediction performance across follow-up time intervals

To examine the temporal robustness of the proposed models, we conducted a stratified evaluation of prediction performance across different follow-up horizons. The dataset was first split into training, validation, and test sets using an age-and sex-stratified partition (80% / 10% / 10%), ensuring comparable demographic distributions across all subsets. Model training and early stopping were performed using the full training and validation sets, while stratified analyses were conducted exclusively at the testing stage.

Specifically, individuals in the test set were grouped according to the follow-up duration from baseline to event occurrence or censoring into three intervals: 0–5 years, 5–10 years, and >10 years. For each interval, we computed the macro-averaged area under the ROC curve (AUC) and balanced accuracy(ACC), as summarized in Fig.4c.

#### Validation of model on GNPC dataset

To assess cross-dataset generalization, we performed an external validation on GNPC dataset. We first identified 2,137 proteins shared between UK Biobank and GNPC. Using feature–label correlations computed on the UKB training set, we selected the top 20% most correlated proteins and applied the same feature set consistently across all methods.

Models were trained and validated on UKB, and then externally tested on GNPC for 19 clinical outcomes. Disease definitions in GNPC were aligned to UKB diagnostic codes through a one-to-many mapping to ensure semantic consistency, covering: stroke, transient ischemic attack (TIA), traumatic brain injury (TBI), Alzheimer’s disease (AD), frontotemporal dementia (FTD), Parkinson’s disease (PD), amyotrophic lateral sclerosis (ALS), mild cognitive impairment / subjective cognitive impairment (MCI/SCI), cancer, diabetes, congestive heart failure (CHF), chronic obstructive pulmonary disease (COPD), myocardial infarction (MI), atrial fibrillation (AF), angina, hyperlipidemia, hypertension, depression, and anxiety.

For fair comparison, all baselines used identical random seeds, data partitioning, and normalization protocols. To reduce bias from sparse outcomes or unstable predictions, we reported summary statistics only on a filtered set of diseases, retaining tasks for which at least one method achieved AUC ≥ 0.6 on GNPC. Thus, all reported results correspond to stable and directly comparable external-test tasks.

#### Validation of autism and schizophrenia prediction on independent sites

Beyond external dataset transfer, we evaluated independent-site generalization on two multi-site neuroimaging cohorts: ABIDE and the Zhangjiang Imaging Brain Bank (ZIB).

For ABIDE, we adopted a leave-one-site-out (LOSO) protocol for autism spectrum disorder (ASD) prediction. Each subject’s brain functional networks was represented by approximately 19,000 functional connectivities from the 200-region Schaefer parcellation^35^. Feature selection was performed using label correlations computed on the training sites, retaining the top 10% most correlated features. ABIDE contains 34 acquisition sites; following common practice, we included only sites with more than 25 subjects. To ensure stability, we further reported results on a filtered set of sites where at least one method achieved AUC ≥ 0.6.

For ZIB, we conducted a similar LOSO evaluation for schizophrenia prediction. The dataset comprises 31 brain volume IDP features (via FreeSurfer) collected across 17 sites. We excluded one site because it contained only a single class, leaving 16 sites for the final LOSO analysis. Again, final statistics were aggregated only over sites satisfying the AUC ≥ 0.6 criterion.

Across ABIDE and ZIB experiments, all methods shared the same random seeds, data splitting, and normalization pipeline, so observed differences can be attributed to modeling choices rather than preprocessing variation.

## Statistical analysis

For large-scale disease prediction in the UK Biobank, model performance was summarized using the area under the receiver operating characteristic curve (AUC) and balanced accuracy (ACC). Within each modality, performance differences between OmicFormer and competing baselines were evaluated across diseases using paired two-sided tests, with each disease prediction task treated as one paired observation. For continuous phenotype prediction, Pearson’s correlation coefficient and mean squared error (MSE) were computed for each trait. Statistical significance of performance differences between methods was assessed across traits using paired two-sided tests, with each trait treated as one paired observation. For MSE, lower values indicate better performance, and relative improvements were defordingly.

To assess the benefit of multi-task learning, single-task and multi-task models were compared within each modality using paired two-sided tests across disease tasks. Each disease served as one paired observation, and AUC and ACC were analyzed separately. This analysis was intended to determine whether shared representation learning conferred systematic advantages over independently trained task-specific models.

For external validation on the GNPC cohort, models were trained on the UK Biobank and evaluated on an independent GNPC proteomics cohort. Disease-specific AUC and ACC were first computed for each disease. Statistical comparisons between methods were then restricted to diseases for which at least one of the compared models achieved AUC ≥ 0.6, thereby focusing on tasks with meaningful discriminative signal. Significance was assessed separately for each retained disease using paired bootstrap tests with 5,000 random resamples with replacement at the sample level. In each bootstrap replicate, identical resampled subject indices were used for both methods to preserve the paired design.

For imbalanced classification tasks, bootstrap resampling was stratified by the true label to maintain class proportions.

To improve interpretability across tasks, performance gains were summarized differently for classification and continuous prediction settings. For classification metrics in which higher values indicate better performance (for example, AUC and ACC), improvements were expressed as relative improvements, defined as (*m*_new_ − *m*_base_)/(1 − *m*_base_). This quantifies the fraction of the remaining gap to the theoretical optimum that is closed by the new model. For continuous phenotype prediction, improvements were reported as conventional relative improvements. For correlation, this was defined as (*m*_new_ − *m*_base_)/∣ *m*_base_ ∣, whereas for MSE, where lower values indicate better performance, relative improvement was defined as (*m*_base_
− *m*_new_)/*m*_base_.

For independent multi-center neuroimaging validation on ABIDE and ZIB, leave-one-site-out evaluation was performed such that each sample was predicted exactly once when its acquisition site was held out for testing. To quantify overall generalization under site distribution shift, all held-out predictions across sites were pooled, and AUC and ACC were computed on the aggregated out-of-site prediction set. Statistical differences between methods were evaluated using paired bootstrap tests with 5,000 random resamples with replacement at the sample level, again using identical resampled indices for the compared methods. Because each subject contributed exactly one out-of-site prediction, this procedure provided a paired assessment of model differences over the full independent validation cohort. For classification metrics, stratified bootstrap resampling by true label was applied to improve robustness under class imbalance. For ZIB site-level analysis, statistical differences within each site were further evaluated using paired bootstrap permutation t-tests with 1,000 random resamples and paired permutations. Within each resample, identical subject indices were used for the compared methods to preserve the paired design.

Unless otherwise specified, all statistical tests were two-sided. In boxplots and summary figures, each point represents one disease task or one continuous trait. Error bars denote 95% confidence intervals. For task-level summaries, confidence intervals were calculated as 1.96 × standard error across tasks or traits; for bootstrap-based external and independent validation analyses, confidence intervals were estimated from the empirical distribution.

